# Data-driven Prediction of Fifteen-Year All-Cause Mortality among 2.3 Million Individuals in the VA

**DOI:** 10.64898/2026.06.29.26356460

**Authors:** Sayera Dhaubhadel, Judith D. Cohn, Tanmoy Bhattacharya, Ruy M. Ribeiro, Kumkum Ganguly, Nicolas Hengartner, Janet P. Tate, Lauren Costa, Yuk-Lam Ho, Kelly Cho, Jean C. Beckham, Nathan A. Kimbrel, Amy C. Justice, Benjamin H. McMahon

## Abstract

We present a data-driven framework to predict 15-year all-cause mortality using outpatient administrative records for 2.3 million Veterans in the largest integrated U.S. healthcare system. Rather than relying on predefined clinical phenotypes, we used the 1,000 most common outpatient medical codes from each of three data types/modalities – ICD-9 (Dx), Current Procedural Terminology (CPT), and prescription drugs (Rx), encoded as binary features. Using these features, we trained three machine learning (ML) algorithms (logistic regression with lasso, random forest, and a 3-layered feed-forward neural network) to predict 15-year mortality risk. The features were also mapped to variables for the widely used Charlson Comorbidity Index (CCI), Elixhauser, and Veterans Aging Cohort Study (VACS) indices, refitted for 15-year mortality prediction, for baseline comparison. All our models significantly outperformed the widely used CCI, Elixhauser, and VACS indices, with C-statistics ranging from 0.82 to 0.84 versus 0.739–0.804 for the baselines. Relative improvements in C-statistics of our approach over the baselines were consistent across different subgroups (age groups of <65 years, those 65+years, Blacks, Hispanics, etc.) Our approach enabled the identification of high-impact predictors with clinical grounding, without requiring hand-curated phenotypes. Cardiovascular diseases and mental health diagnoses/treatments emerged as leading long-term mortality indicators. Using unsupervised ML techniques including PCA and K-means clustering, we associated interpretable patterns and complex interactions between diagnoses and treatments, highlighting comorbidities, disease trajectories, and healthcare utilization patterns. The ability to achieve the predictive performance and algorithmically detect such relationships purely from outpatient data supports the scalability and broad applicability of our framework. This framework not only improves mortality risk stratification over existing clinical indices, but also enables better understanding of how medical codes, regardless of category, interact to predict long-term outcomes.

## Introduction

Accurate and reliable estimation of all-cause mortality risk is foundational for maximizing health care value. Among others, applications include precision population-health management, risk-adjusted quality benchmarking, and downstream causal and comparative effectiveness analyses. Because real-world observational electronic medical record (EMR) data are plagued by confounders, sporadic measurement errors with variable standards, informative censoring, and selection bias, naïve statistical adjustment is rarely sufficient. Randomized controlled trials (RCTs) minimize such confounding effects but are expensive, slow, and often ethically or logistically infeasible. Consequently, modern comparative-effectiveness frameworks integrate advanced prediction methodologies with high-dimensional observational data to complement RCT evidence^1–4^.

Long-term medical risk prediction can use inpatient and/or outpatient information. Inpatient records, often enriched in acute physiological conditions, excel at short-horizon predictions, whereas longitudinal outpatient utilization patterns encode chronic multi-comorbidity trajectories and health-seeking behaviors that are critical for decade-scale forecasts. When granular EMRs are unavailable, composite indices derived from routinely coded comorbidities, like the CCI, Elixhauser, and VACS indices — provide compact proxies for baseline disease burden^5–8^; recent work has re-validated and re-calibrated the CCI and VACS in contemporary populations, underscoring its enduring utility^9, 10^.

The advent of scalable ML pipelines has enabled modeling of multi-modal EMR data (diagnoses, procedures, prescriptions, clinical text and imaging) for cohorts numbering in the millions^11, 12^. Yet systematic evaluations reveal that cross-site and cross-population generalizability often degrades sharply, even for highly expressive models, if training data lack population, geographical, and institutional diversity, stressing the need for external validation and domain adaptation^13^. Furthermore, ablation studies demonstrate that feature engineering choices (e.g., choice of variables, longitudinal aggregation windows, hierarchical code grouping, and temporal decay functions) can account for more performance variance than the specific learning algorithm itself, implying that transparent representation learning and rigorous model selection should precede complexity escalation^14, 15^.

The Veterans Health Administration (VHA) corporate data warehouse (CDW)^16^ constitutes one of the world’s largest integrated longitudinal EMR repositories (over 25 million Veterans, since 2000). Leveraging a 20-year window of the CDW, we encode the 1,000 most prevalent outpatient codes for each of the Dx, CPT, and Rx domains into sparse binary predictors from a five-year observation window, and train automatically-regularized interval prediction models to predict subsequent 15-year all-cause mortality. Our objectives in this study are three-fold: (i) to derive a parsimonious predictive model that operates solely on outpatient data without the need for hand-curated phenotypes and compare it to the standard baselines, (ii) to algorithmically identify a wide array of outpatient descriptors with clinical grounding that describe contributions to our mortality predictions, and (iii) to characterize the comorbidities and trajectories of medical conditions using outpatient descriptors.

In this study, we demonstrate the sufficiency of high-dimensional, minimally curated outpatient medical codes in predicting long term mortality by utilizing multiple ML algorithms to develop models and comparing them to widely used clinical indices. Moreover, we characterize patterns of comorbidity and complex dependencies/interactions among Dx, CPT, Rx, and demographic variables, algorithmically, when different combinations of such variable classes are included as predictors. Comparing results from different models to understand the drivers of prediction (e.g. age vs. specific conditions, Dx vs. Rx, or linear vs. nonlinear models, we uncover the role of surrogate drivers/variables in the absence of the real ones. This work aims to serve as a proof-of-concept upon which other studies can be conducted. Our study can serve as a framework for data-driven prediction as well as characterization of comorbidities and drivers of mortality at scale that can be easily ported across different populations and healthcare systems.

## Results

The results of our study are described in three sections. First, we describe our model and compare its accuracy to three commonly used clinical indices. Second, we characterize the predictor variables in detail to understand their associations with each other and the outcome. Finally, we analyze in detail the tradeoffs that occur with the inclusion/exclusion of age, demographics, and other classes of variables.

We note a wide variety of selection biases and correlations in our study of Veterans in the US Department of Veterans Affairs (VA), which is predominantly white males. Biases include women being ten years younger than men, while correlations include that the median age of individuals with schizophrenia (55 years) is 16 years younger than the median age of individuals diagnosed with a heart attack. We present a sample of these correlations in Table S1.

## Predictive Modeling Accuracy

Fig. 1 presents the C-statistic (or AUROC) scores for predicting 15-year mortality among the *∼*1.15M individuals in our test dataset using three prediction methodologies applied to nine sets of predictor variables. The predictive performance of the models on the training dataset (not shown) were comparable to the test dataset. In comparison to three standard clinical indices used for all-cause mortality – CCI, Elixhauser, and VACS indices, our models using high dimensional predictors achieved 5–16% higher C-statistic.

**Figure 1.**
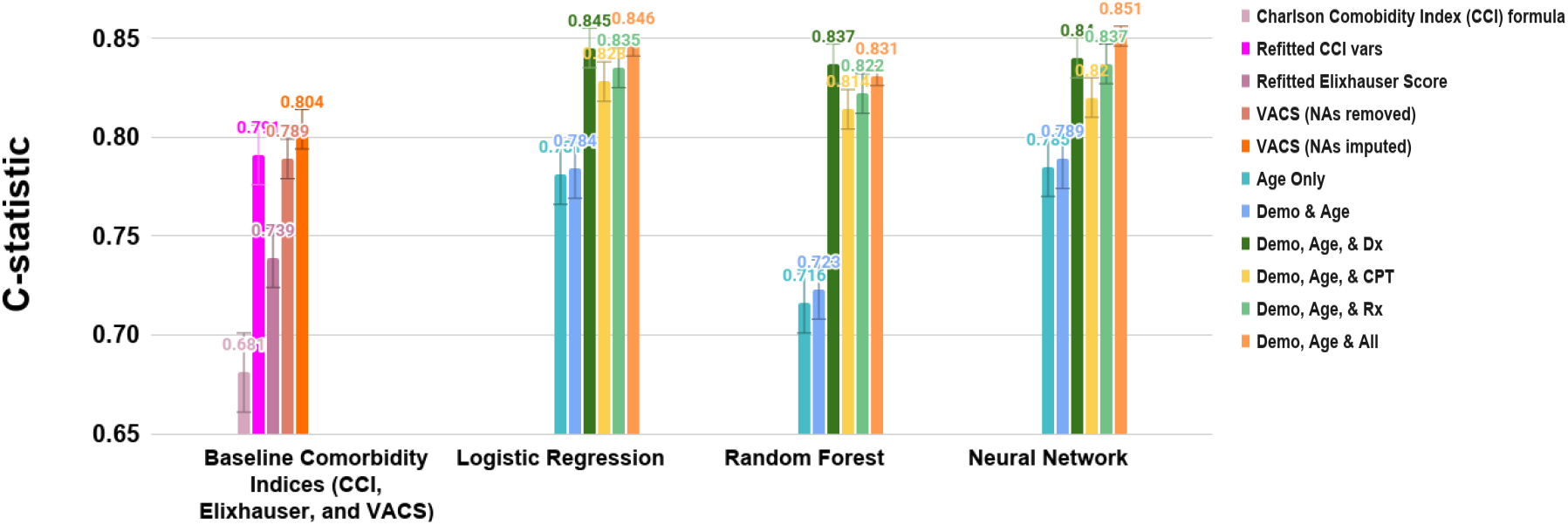
C-statistics to predict 15-year all-cause mortality on the test set (*∼*50%) of the overall 2.3 million veterans using four modeling methodologies and trained on nine sets of predictor variables. Patient data were split into age-matched data sets as train and test sets. The logistic regressions were implemented together with lasso model selection for the individual data modalities, then only model selected variables were used in the combined regression analysis.

The C-statistic for logistic regression with lasso model selection ranges from 0.78 when using age alone (presented to the model as age, age^2^, age^3^, and age^4^) to 0.84 when all variables, including age, demographics, diagnoses (Dx), procedures (CPT), and generic drug names (Rx) are used for predictions. The biggest increase in predictive performance occurs when Dx codes are added, but including information on either CPT or Rx codes is almost as valuable, with Rx codes leading to slightly more accurate predictions than CPT. Combining Dx, CPT, and Rx variables offers only slight improvement in accuracy of mortality prediction over using individual sets of codes alone. However, we observe smaller variation in predictive performance with different predictive modeling algorithms. Since the logistic regression was easiest to interpret with predictive performance comparable to the best, we utilize this calculation, with all variables included, for the rest of our analyses, unless noted otherwise. Fig. S1 show good calibration of our logistic regression models across three important subgroups: age, gender, and race-ethnicity.

Table S1 shows a detailed characterization of the 2,322,813 individuals in our study, providing better insight into our cohort design and predictive performance. Our dataset is balanced in terms of the two possible outcomes with 47.6% of patients linked to a recorded death. The median age of all patients is 60.7 years at the time of prediction, with the first and third quartiles at 47.2 and 76.5 years respectively. People who died in the follow-up period are about 10 years older than people who were alive after the follow-up period. Females, Blacks, and Hispanics represent only 7.9%, 16.1%, and 4.7% respectively, of the study population, and are younger (median age of 50.9, 55.0, and years, respectively) than the overall patient average for the data set. Another metric for clearer characterization of this data set is provided in columns three–five, which shows that in changing from the oldest to youngest quartile, the percentage of females changes from 1.6% to 18.8%, while the number of Blacks increases from 7.6% to 27.8%. Further details are discussed in Section SI-1.

The observation that similar prediction accuracy is possible with each modality of data (Dx, CPT, and Rx) suggests that there is significant redundancy in information across the three modalities of medical codes, with the Dx codes providing the most complete information. That treatments alone (CPT and Rx codes) can be used to effectively predict mortality suggests that the models may be using the presence of these codes not as mitigation of disease, but as redundant indicators of disease or indicators of disease severity. The similar performance of the model utilizing all data classes when compared to that with single data class provides further indication that 1,000 variables provides sufficient flexibility and information to make accurate predictions of 15-year mortality. The robustness of this result across several prediction methodologies and minimal change in predictive performance when evaluated on the test dataset vs. the training dataset ∼50-50 split) suggests we may have saturated our ability to predict mortality 15 years in the future with these cross-sectional data.

### Characterization of the Predictor Variables

We characterize the relationship between the medical codes (Dx, CPT, and Rx) as well as the comorbidities and disease trajectories, based on the cosine distance (or dissimilarity) of the medical codes and the patient profile-based correlations among the Dx, CPT, and Rx codes.

### Causes of Mortality

Fig. 2 presents the first two principal components (PCs) from principal component analysis (PCA)^17, 18^ applied to the cosine distance (or dissimilarity) matrix^19^ of the medical codes obtained from the patient profile. Each point on the plot represents a Dx, CPT, or Rx code among our predictor variables. The two PCs accounted for 59% and 35% of the variance, respectively. Fig. 2a, showing only the 1000 Dx codes and color-coded by the International Classification of Diseases (ICD)-9 categories, disperses the codes roughly into arcs, corresponding loosely to the major ICD-9 categories of disease. cardiovascular diseases (CVD), diabetes, mental health, and dental codes appear as the most prominent predictors (see inset) among the different ICD-9 categories.

**Figure 2.**
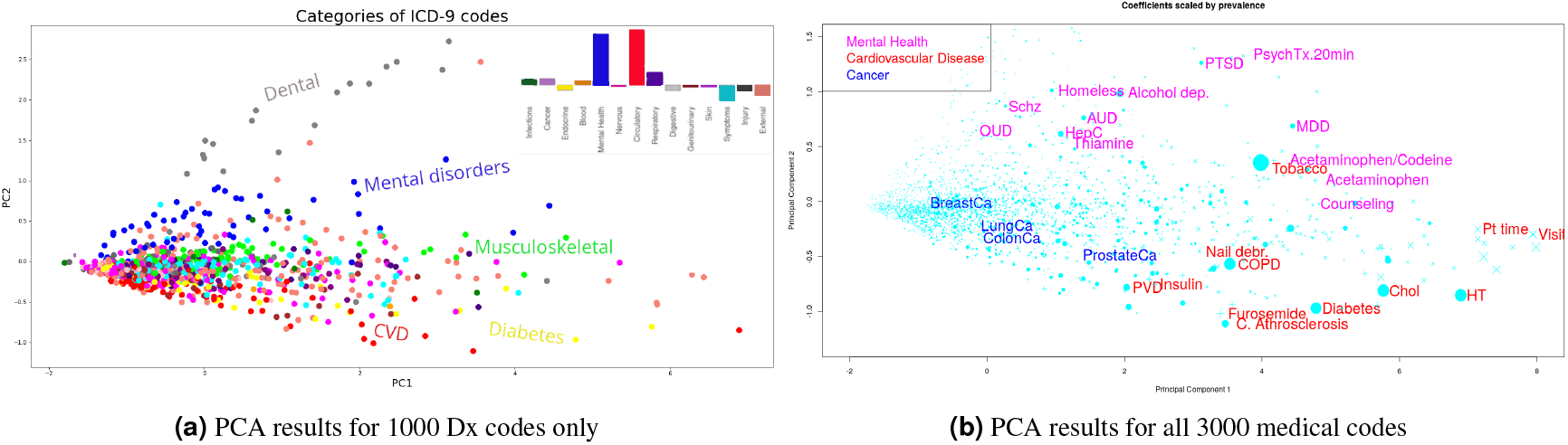
Plot of first two principal components from PCA applied to the cosine distance (dissimilarity) matrix of the 3000 medical codes of our study. **(Left)** shows only the 1000 Dx codes, color-coded based on ICD-9 categories. The bar plot (inset) shows the sum of the `coefficient` *×* `prevalence` for individual Dx codes within each ICD-9 category. **(Right)** shows all 3000 codes color-coded as representative of mental health, CVD, and cancer related codes, with symbol size proportional to `coefficient` *×* `prevalence`.

Within this analysis, we provide estimates of the population level association of individual ICD-9 category of disease to long term all-cause mortality, shown in the bar plot of Fig. 2a. These associations are estimated by taking the sum of the product of coefficient and prevalence for individual codes within the category. This plot suggests an important finding that the mental health category is the second most important category of predictors for predicting long-term mortality. This is further substantiated by the median age at death for patients with mental health conditions which is 65 years in comparison to the median age at death for patients with cardiovascular conditions which is 75 years, as shown in Table S1.

In Fig. 2b, which shows all 3000 medical codes in our study, we observe that the first PC is identifiable with the prevalence of the code, while the second component defines a continuum of patient types, with CVD and diabetes at one extreme, and mental and dental codes at the other. An interesting result of the PCA analysis, obtained with a pure data-driven approach, is the spectrum of comorbidities afflicting individuals, showing how common disease, procedures, and treatments as well as behaviors are intertwined and are either predictive (with positive coefficient) or protective (with negative coefficient) of mortality.

We can bring together the predictive modeling, above, with analyses of the cosine distance of codes across patients. Figure S3 shows the plot of the first two PCs of this matrix, where the symbol-size corresponding to each code is proportional to the prevalence of the code times the coefficient of the code from the combined logistic regression model. Symbols are colored green to indicate negative coefficients (protective) and red when positive (predictive of mortality). In this figure, we annotate selected symbols, in order to understand how our method associates multi-modal data for the particular examples of cardiovascular disease (towards the bottom of the Fig. S3 and mental health-related issues (towards the top of Fig. S3). One of the largest symbols on the plot belongs to tobacco use disorder, which is predictive of mortality in both types of patients, appearing close to the center of the figure. Figure S3 shows the three types of medical codes used in our study nicely interspersed; related codes from different categories are located close together in the plot, regardless of their original categorization defined in the ICD (Dx), CPT, and Rx handbook. Detailed analyses of the comorbidities and disease trajectories for cardiovascular diseases, and mental health disorders are presented in Section SI-2. These results show promising utility of our data-driven framework that can be applied to other populations to understand and characterize the standard of healthcare in the population.

### Disease Clusters and Multi-modal Descriptors

In Fig. 3a, we present the results of k-means clustering applied to 750 medical codes, 250 from each of the most common Dx, CPT, and Rx variables, based on their occurrence among patients in our study. The clusters are then organized and visualized using a minimum spanning tree algorithm to produce a graphical (tree) structure. This clusters 750 medical codes into 150 groups of medical conditions based on the similarity of the profile of patients assigned each code. The tree shown in Fig. 3a has 1/4 of the common codes at the center, and 15 identifiable groups of codes radiating out from this center. The more common codes are towards the center, and less common, but often more serious conditions, towards the edges. This clustering approach provides lists of 10–100 codes in identifiable categories of medical conditions (heart failure, depression, chronic pain, etc.) which need to be considered in the context of each other.

**Figure 3.**
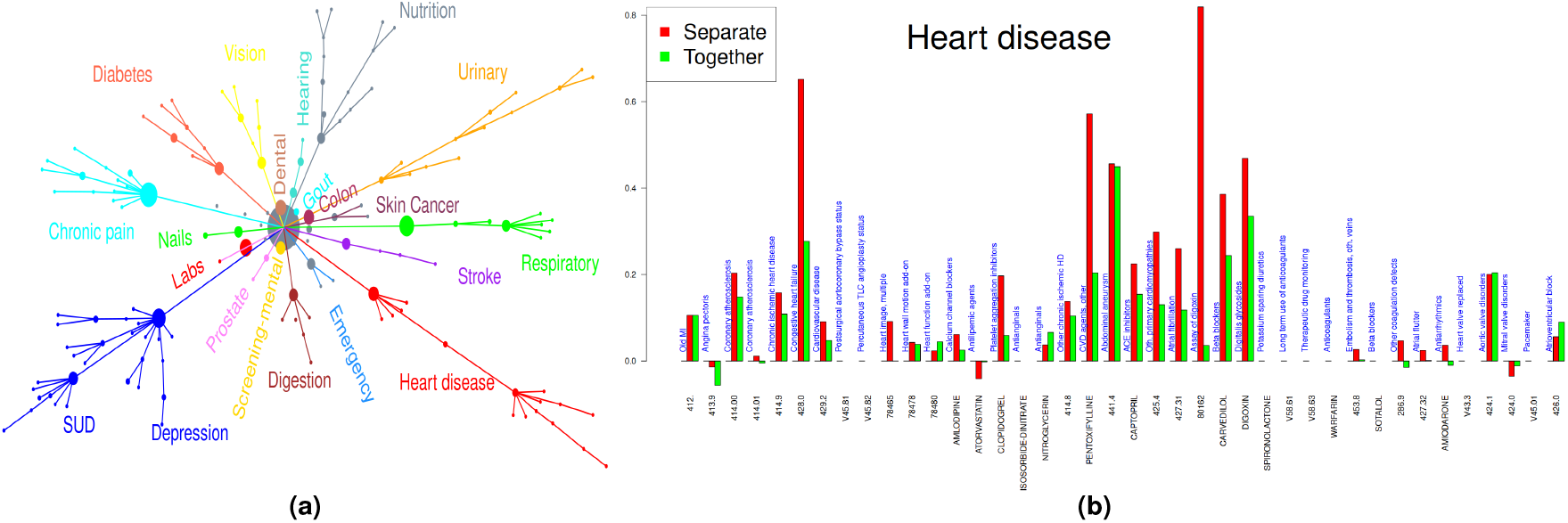
**(Left)** The 250 most common of each code type (Dx, CPT, and Rx) were clustered based on their overlapping occurrence. Clusters were labeled based on membership. The area of the circle at each node is proportional to the number of codes comprising the cluster. The center (gray) node is comprised of *∼*200 commonly occurring codes, such as those to the right side of S3. **(Right)** Coefficients from the logistic regression models for the medical codes (*heart disease*) cluster from Fig. 3a. Two bars are shown for each code. Red bars, typically longer, show the model coefficients for the Dx, CPT, or Rx codes from the three models where single type of codes are used as predictors. Green bars show the coefficients from the single model when all three types of codes are combined as predictors. Positive coefficients extend above and negative below the horizontal line. The codes are ordered in groups from the center of the graph, sorted by type of code within each group.

Fig. 3b shows a representative example of one of the important clusters (*heart disease*) identified using the k-means clustering and the minimum spanning tree algorithms, with details explained in the caption. Evaluation of the model coefficients of treatments (CPT and Rx) with and without diagnoses (Dx), or *vice-versa*, suggests that treatments can be redundant information with diagnoses. An example of this in Fig. 3b is provided in the set of codes for *Congestive heart failure, Digoxin*, and *Assay for digoxin*. When only a single domain of codes are used, each of these carries a relatively high risk; when used in the combined model, the risk is apportioned among the three in a way that may convey additional information about diagnosis, treatment, and disease severity. On the contrary, there may be diagnosis codes for which there is no specific treatment, such as *Carotid artery occlusion (433*.*10)* or *Aortic aneurysm (441*.*4)*, where the coefficients are within 1% in the models with and without treatments included. There are numerous instances of Dx codes or treatments that are important primarily because they are indicative of patient behaviors, such as level of preventative care obtained, or health care utilization habits (eg. vaccination). In this case, coefficients will differ because the different models provide different basis sets on which to project these behaviors. A more detailed analyses of coefficients within clusters is presented in SI-3 with examples of other important clusters for diabetes, substance use disorder, and depression shown in Figure S4.

### Interplay of Age and Classes of Medical Codes in Predicting Mortality

In Fig. 4, we investigate the association of our predictor variables with mortality when applied in different combinations with age. Fig. 4a shows the baseline (attributed to age) probability of mortality in 15 years as a function of age at the time of prediction, conditioned upon survival until the age of prediction, from three models trained with – i) age only (solid curve), ii) age, demographics, and the 1000 Dx codes (dashed curve), and iii) age, demographics and all 3000 medical codes (dotted curve). The most striking difference between the baseline probabilities of mortality in the three models shown in Fig. 4a is the *∼*80% reduction from the age-only model when all three classes of data were included, compared to a *∼*20% reduction when only one domain of data (Dx: shown, or Rx, CPT: not shown). Evidently, when age-alone predicts mortality (solid curve) it serves as a surrogate for other medical conditions. Hence, in the absence of specific diagnosis or treatment codes, age can reasonably accurately predict of mortality, with a C-statistic of 0.781. The colored points show the average age vs. average mortality for selected rows from Table S1 graphically, so that fractional mortality of cohorts can be easily compared to baseline calculations of age-dependent 15-year mortality probabilities. They are further discussed below.

**Figure 4.**
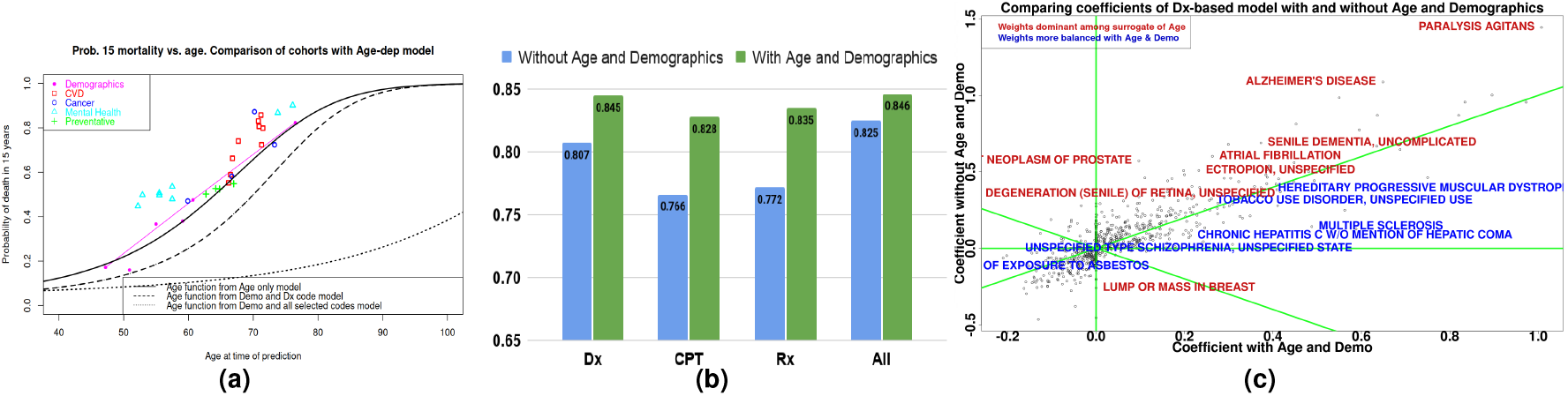
**(a)** Baseline probability of dying within 15 years conditional upon survival at the time of prediction from several models, compared to cohorts of patients defined in Table S1. The thick solid line results from a logistic regression model trained with only age, age^2^, age^3^, and age^4^. The dashed and dotted lines reflect the baseline (considering only age) conditional probability of 15-year mortality estimated from age alone in a model trained with the Dx codes, and with all Dx, CPT, and Rx codes, respectively. Symbols are placed at the median age and 15-year survival fraction for each cohort defined by the rows in Table S1, color-coded by category. **(b)** C-statistic scores for the four combinations of medical codes are used as predictors with and without age and demographics **(c)** Coefficients of Dx-codes-based logistic regression model with and without age and demographics.

Another way to understand the role of age in different models is in Fig. 4b, which shows the C-statistic scores when the four combinations of medical codes are used as predictors with and without the inclusion of age and demographic. When a single domain of medical codes (Dx, CPT, Rx) are combined with age, the C-statistic improves by 0.04-0.06, but the most of the risk is still captured by age (dashed line in Figure 4a), modulated by the medical codes. When using all three domains of codes, however, adding age only increases the C-statistic by 0.021, consistent with age making only a minor contribution (dotted line in Figure 4a). Fig. 4c is a plot the coefficients of Dx-codes-based logistic regression models with and without age and demographics, showing which codes change the most when age is eliminated from the model. As expected, these are conditions with a strong age-dependence, including Alzheimer’s, senile dementia, paralysis agitans (Parkinson’s), ectropion, and prostate cancer, shown in red).

Finally, we return to the colored points in Fig. 4a, which show the average probability of 15-year mortality vs. average age for representative medical conditions presented in Table S1. These provide the average elevated risk for each of the medical conditions (in a univariate setting) in comparison to the baseline age dependent risk curves. For example, a diagnosis of opioid use disorder (median age 52.9 years and 49.8% mortality) approximately doubles the estimated probability of 15-year mortality, while heart failure (median 71.2 years and 85.8% mortality) reflects a much smaller change relative to the baseline of 70%. This helps us to understand the perhaps unexpected result shown in the inset of Figure 2a, that the total population attributable risk (`sum of coefficient` × `prevalence`) for mental health codes is nearly equal to that of CVD. Because mental health conditions afflict younger Veterans than CVD, a larger risk is required to effect mortality, while cases where mental health conditions contributing to a CVD death will not be identified as a proximal cause.

Table S2 shows the variation C-statistics (AUROC) and AUPRC scores of the Age-Demo-Dx model, when tested on with certain sets of Dx codes randomized. The top panel shows the scores on the entire test data set and while the bottom shows the scores on a subset with lower probability of mortality (logit score *<* -1), i.e. individuals whose risk are harder to predict. Examination of the drop in the performance scores further substantiates this observation of the importance of mental health related codes. Modest reduction in scores is observed when mental-health and/or CVD related codes are randomized, i.e. the effect of that specific set of variables is nullified. We hypothesize that the modest reduction is due to correlations between one set of codes with other medical condition that serve as parallel predictors. Moreover, with a C-statistic score for predicting mortality almost reaching to a saturation point, even minimal reductions/improvements are significant. This argument is corroborated the significant drop in C-statistic score when tested on a subset of (*∼* 50% of all) samples with lower predicted probability of mortality, i.e. samples that are harder to predict.

## Discussion

Our study on the VA EMRs is exceptionally rich in the number individuals, duration of follow up, and the breadth of available information. We use these data as a testbed for evaluating the interplay of prediction methodologies and input data types on 15-year mortality prediction on a balanced data set with over one million individuals each as cases and controls.

### Model Accuracy and Logic of Prediction

Using minimally curated high dimensional outpatient medical codes, we were able to develop robust and well-calibrated models to predict 15-year all-cause mortality with high accuracy. Our models performed much better (0.85 vs. 0.739–0.791) than existing indices including CCI^5, 20^, Elixhauser^6^, and VACS^7, 8^. The statistical significance of a binary prediction evaluated across one million samples in a balanced data set is 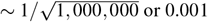, while the importance of such a difference depends on the details of any particular difference.

Following common practice, we used the C-statistic, or area under the ROC curve (AUROC), on a single prospective cohort as a metric of model performance across prediction variables and methodologies. Cook^21^ discusses in detail the caveats of using of C-statistics to compare models, including her observation that 0.83 is the maximum possible C-statistic for a well-calibrated model (see Fig. S1) predicting for a uniform probability distribution, (similar to our case, data not shown). We also evaluated an Age-only model that suggests 0.781 as a baseline model accuracy for comparison, provide calibration curves in important demographic subgroups (Fig. S1) and AUPRC and randomization of patients across subsets of variables (Table S2) for the additional insights these metrics provide.

After noting that logistic regression with LASSO model selection performs nearly as well as our best-performing model, neural networks, we exploited the interpretability advantage of the linear model to dissect the contributions of age, diagnoses, procedures, and medication codes to model performance. In doing this, we find both conceptual and practical difficulties in apportioning all-cause mortality risk among these groups of codes. Our observations are consistent with previous studies, including our previous work predicting suicide-related outcomes among millions of Veterans^22^, on the importance of patient behaviors, missingness of data, and interactions with age in predicting medical outcomes using thousands of predictor variables. By choosing the well-defined outcome of all-cause mortality we minimize an important source of confounding in our observational study.

While we focused our interpretability efforts on our linear model, our knowledge of the complex non-linear interactions between the predictor variables such as age, diagnoses, and treatments suggest non-linear models are required to accurately reflect reality^23, 24^. While our neural network model does not accurately identify important interaction terms amongst predictor variables,^25^ it is reassuring to see that our best-performing model (C-statistic of 0.851) was the neural network on all of the input variables, and that it was close to the maximum expected accuracy when predicting on our probability distribution. Our approach of analyzing both models, where the linear model provides greater understanding relationships between the predictor variables, while the non-linear method can be used to maximize prediction accuracy.

### Drivers of 15-year Mortality

Even for acute mortality prediction, the cause of death can be multifactorial. For 15-year mortality, this problem is exacerbated to the extent that age-alone provides a competitive baseline prediction model. In keeping with our desire to identify chronic diseases and healthcare utilization practices predictive of mortality, we chose to use outpatient codes to predict 15-year mortality rather than the inpatient codes and labs used for the 5-year mortality predictions of the comparator indices. We also exploited the apportionment of risk intrinsic to the coefficients of logistic regression using binary variables to associate both effect size (*coe f f icient*) and population attributable risk (*coe f f icient × prevalence*) to both individual and groups of medical codes.

Our Dx and demographics model shown in Figure 2a showed that CVD and mental health conditions were similar and predominant drivers of 15-year mortality. While CVD provides many of the major causes of death listed by the CDC^26^ (heart disease, cancer, unintentional accidents, chronic lower respiratory disease, stroke) the same cannot be said of mental health conditions. While certain psychiatric conditions such as schizophrenia are related to early mortality^27–29^, examination of our Figure 4a and Table S1 suggests an important distinction is that mental health conditions most strongly impacts patients twenty years earlier than CVD, when the overall mortality rate is much lower. Thus, mental health conditions are frequently listed as **contributing** causes of mortality^30^, and that mental health condition can improve mortality risk prediction^10^. A second notable feature in the bar plot of Fig. 2a is the relatively low importance ascribed to neoplasms as a category. This was observed despite the relatively high mortality rates identified in Table S1. This may be explained partly by the advanced age at which many cancer deaths occur and their correlation with CVD, and partly because cancer often occurs within a few years of initial diagnosis, and so is often invisible to our 15-year study design.

An important limitation of linear models arises from patterns of co-occurrence of medical codes, which can arise for many reasons, including disease progression, treatments associated with diagnoses, healthcare utilization patterns and other patterns of patient and provider behaviors. Because future attempts to account for and exploit these patterns will require an understanding of the source of correlations, we characterized them in several ways, including PCA of the co-occurrence matrix clustering of codes and observing the sensitivity of model coefficients to changes inclusion of other variables such as age, diagnoses, procedures, and medications. These analyses are described in the Results section and supplementary information, we hope they will provide the reader with both conceptual and practical value in disentangling our high-dimensional mortality calculation.

### Limitations and Future Directions

Our study has several important limitations. Compared to the general population, Veterans in care are less likely to be Female, are means tested, and may have distinctive occupational exposures. Hence, our findings in this population may not translate directly to the general population and details of the associations must be interpreted with caution. Likewise, the risk calibration for particular subgroups such as Female, Black, or Hispanic (Fig. S1) needs further investigation and improvement. Having a model that is right for the right reasons should improve the generalizability, as described in Justice *et al*.^31^, and a more rigorous test in a different population as done in Jensen et al.^32^ is necessary to fully assess the utility of our approach.

We have not explored the potential advantages of temporal modeling with more sophisticated ML models like RNNs or Transformers in this study. We suspect that the gains of such temporal models utilizing detailed sequence of events would be more pronounced for modeling of more acute conditions including survival in a short time. Use of longitudinal data may still result in modest performance improvement, particularly for more individuals with serious medical conditions.

We have shown that high-dimensional data sets with non-linear predictive models greatly outperform existing clinical indicies (> 0.05 increase in C-statistic), yet the inherent co-linearity and inter-dependencies among predictor variables make it highly unlikely that the model itself will be directly useful in other healthcare systems or even several years into the future, when for example ICD-9 codes are no longer used. Nevertheless, our data-driven approach can be easily ported and applied to other healthcare systems in the presence of sufficient data. We expect that the information and techniques presented here can be used to create similar models, even in situations with far less training data, by using techniques such as careful variable definitions and ensemble transfer learning, that we used to predict suicide amongst millions of Veterans^22^.

## Conclusion

We have demonstrated a systematic data-driven approach to develop and validate stable predictive models for 15-year all-cause mortality using thousands of outpatient medical codes as predictors in a study involving over 2.3 million patients. Our models outperformed existing clinical indices for this question, motivating future work to provide such models with adequate validation and generalizability. Our approach not only demonstrated superior discriminatory power but allowed for characterization of relationships between different medical codes and their association with the outcome when utilized in a completely data-driven manner.

Another important aspect of predictive modeling is disentangling major contributors of outpatient medical diagnoses and treatments to all-cause mortality to ensure that these contributors complement our medical understanding of these codes. Patient profiles and the cosine distance matrix computation of these medical codes across patients provided a purely data-driven process to understand complex relationships between different diagnoses and treatments. The process implicitly mixes different types of medical conditions and treatments because of the high degree of comorbidities and the pervasive nature of patient behaviors being captured by the medical codes. We were able to identify and validate cardiovascular diseases (CVD) and mental health disorders as the major contributors of 15-year mortality.

Fifteen-year all-cause mortality risk after a 5 year observation period of presence or absence of outpatient codes relates chronic medical conditions and healthcare utilization patterns to mortality. This formulation is agnostic about whether a variable is a primary or contributing cause of death, a diagnosis or a treatment, and may be inferring disease severity through co-occurrence patterns of colinear variables.

## Methods

### Cohort Design

Our study used a five year observation period from April 1, 2000 to March 31, 2005, followed by a fifteen year follow up period during which a date of death was ascertained, as shown in Fig. 5.

**Figure 5.**
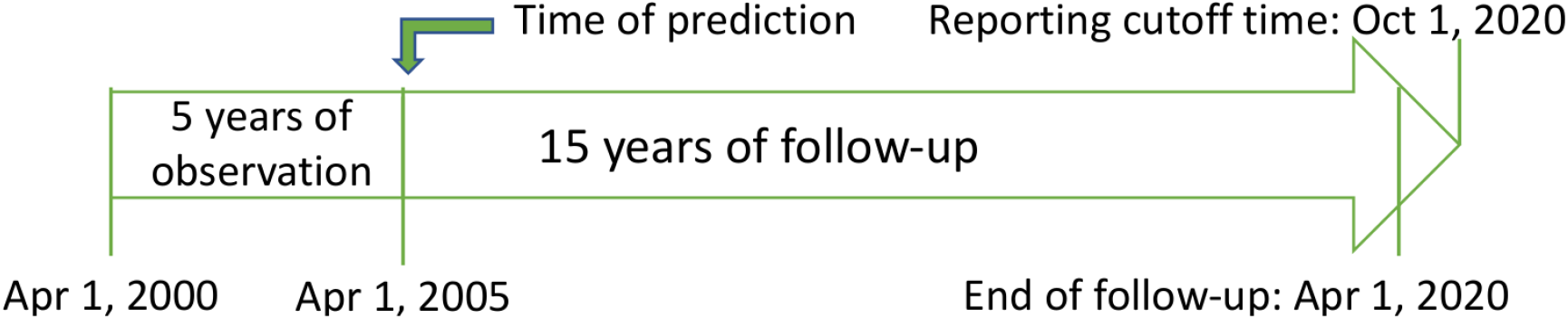
**Study Design**. Individuals were included in our study if they were seen at least once before Oct 01, 2000 and at least once in the observation window (between April 01, 2000 and April 01, 2005). For each of the 2.3 million patients satisfying this criterion, the presence or absence of each of the 1,000 most prevalent Dx, CPT, and Rx codes was recorded over the five year period from April 01, 2000 to April 01, 2005. Individuals with recorded deaths during the 15-year follow-up period from April 1, 2005 to April 01, 2020 were included as cases and assigned an outcome of ‘death’, if that death was reported to the VA records by October 01, 2020. All other individuals without a death record and an encounter with the VHA in the follow-up period were included as controls and assigned an outcome of ‘alive’.

Inclusion criteria for this study were that the patient had a recorded and valid date of birth, was seen at least once before Oct 01, 2000 and at least once more during the observation period (between April 01, 2000 and Mar 31, 2005). This inclusion criteria was chosen to ensure that only ‘active’ patients with enough data are included in the study. Being ‘seen’ was defined as a recorded diagnosis, procedure, pharmacy prescription, or vital sign.

Our study design relies on completeness of death records within the VA data. Comparison of the death records in the VHA with the National Death Index (NDI) obtained from Center for Disease Control (CDC) showed 98.9% concurrence. Relying on this statistic and the attestation of the excellent quality of death data in the VHA in previous studies^8, 33, 34^, individuals without a recorded death in the follow-up period (from April 01, 2005 to October 01, 2020) was presumed to be alive. Individuals with a death record in our follow-up period were included as cases. The controls were chosen from people without a death record on the study end date and had at least one encounter with the VHA in the follow-up period. Individuals who died during the observation window or did not have an encounter with the VHA in the follow-up period were excluded from our study.

### Data Processing

For each of the individuals in the study, information on the 1,000 most common outpatient medical codes for the each of (ICD-9) diagnoses codes (Dx), procedure codes (CPT), and generic drug names, along with age, gender, race, and ethnicity was collected. Each medical code was encoded as present or absent (binary) during the observation period. The data was split equally into age-matched training and test sets with *∼*1.15 M patients in each set, to ensure extensive validation on a wide distribution across all important covariates. Age was used as a continuous variable. Race and ethnicity were assigned according to procedure established in VA technical report^35^ and encoded as Black / African American or not, and Hispanic or not.

### Predictive Model Development and Evaluation

Four different predictive modeling algorithms were used to train models in our analysis and these models were each trained on six different combinations of input data classes—age only, age and demographics (gender, race, ethnicity) only, demographics with Dx codes, demographics with CPT codes, demographics with prescription names, and demographics with all three modalities of codes.

First, we computed the three commonly used clinical risk indices to predict mortality risk on our data. Charlson Comorbidity Index (CCI) was computed with the original point-based scoring as described in the paper^5^. We then use the same predictor variables to fit our own CCI model to the mortality risk outcome of our population for a better comparison. Likewise, we fitted two more models for predicting 15-year mortality risk using Elixhauser^6^ and VACS 2.0 variables^8^.

Second, a logistic regression^36^ model with elastic net^37^ penalty for model selection^38^ was applied using 5-fold cross validation to obtain a trained model with only the ‘most significant’ variables. Logistic regression^39^ is a statistical model that uses a logistic function (a linear model followed by sigmoid function) to model a binary dependent variable and provides a mechanism for applying the techniques of linear regression to classification problems. Three separate elastic net models were trained for the individual modality of the medical codes (ICD-9, CPT, and prescriptions) to get a subset of variables (codes) that were most important in each modality. Then, a combined logistic regression model was trained including demographics and variables selected by the elastic net models trained on the three individual modalities of data.

Third, random forests with 1000 trees and maximum depth of 300 were used on the six combinations of inputs mentioned above. Random forests^40^ are a combination of tree predictors such that each tree depends on the values of a random vector sampled independently and with the same distribution for all trees in the forest. The generalization error of a forest of tree classifiers depends on the strength of the individual trees in the forest and the correlation between them. The number of features to consider when looking for the best split was set to the square root of the total number of features.

Fourth, a simple fully connected (vanilla) neural network containing 3 hidden layers was used. Neural networks^41–43^ are computing systems roughly inspired by the biological neural networks that constitute animal brains. They are based on a collection of connected units or nodes called artificial neurons. Each neuron is essentially a linear weight followed by a non-linearity, and the power of neural networks comes from the combination of hundreds or thousands of such non-linear relationships. ReLU^44, 45^ non-linearity is used in each of the hidden layers and the number of neurons was 512 (1024 for the combined model), 256, and 64 for each of the three hidden layers, from input to output.

In a baseline run, we tested age alone as predictor of death and concluded that including a polynomial in age, age^2^, age^3^, and age^4^, provided a reasonable approximation to an age-dependent probability of death over 15 years. Then, this age polynomial was included as a predictor variable, with a linear coefficient modulating the overall function. However, for random forests and neural networks, we used normalized age in its original form with the belief that these algorithms should be able to pick up the complex non-linear relationship mortality with age.

### Principal component analysis (PCA) and K-means Clustering

We leverage the patient profiles across the 3000 outpatient medical codes to compute the cosine distance between each pair of medical codes in our study. Given the patient profile matrix *A*, the cosine distance matrix *D*^cos^ ∈ ℝ^*n*×*n*^ is given by:

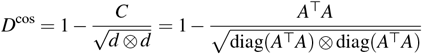

where, *AA*^*T*^ is the raw co-occurrence matrix, *diag*(*A*^*T*^*A*) is the self-co-occurrence (i.e. counts of each item with itself), and *d* ⊗ *d* is outer product of the diagonal, yielding a matrix. The overall matrix calculation returns the cosine similarity where each normalized entry *a*_*i, j*_ represents the cosine similarity between *i* and *j*. The final (1 −*cosine*_*similarity*) gives the cosine distance and represents how dissimilar or distant the two medical codes are.

We then apply PCA ^17^ to this cosine dissimilarity or distance matrix and present plots the first two principal components, accounting for *∼*95% of the overall variance. This unsupervised dimensionality reduction approach allowed the identification and validation of comorbidities and disease trajectories (discussed above in detail) in a completely data-driven approach.

We also apply another unsupervised ML technique – k-means clustering^46^ to the patient profile matrix to extract direct relationships between the individual medical codes. For this, we only use 250 of each (750 total) of the three types of medical codes (Dx, CPT, and Rx). This approach returns discrete clusters of medical codes pertaining to specific medical conditions. Both techniques identify relationships between the different medical codes, and patient behavior without the need to input/encode prior medical knowledge.

## Supporting information

SupplementaryFile_Coeff_Clusters

## Acknowledgements

**Notice:** This manuscript has been authored by Triad National Security, LLC under Contract No. 89233218CNA000001 with the U.S. Department of Energy/National Nuclear Security Administration. The United States Government retains and the publisher, by accepting the article for publication, acknowledges that the United States Government retains a non-exclusive, paid-up, irrevocable, worldwide license to publish or reproduce the published form of this manuscript or allow others to do so, for United States Government purposes. The Government has also granted for itself and others acting on its behalf a nonexclusive, paid-up, irrevocable worldwide license in the code and data, within this manuscript, to reproduce, prepare derivative works, and perform publicly and display publicly, by or on behalf of the Government. NEITHER THE GOVERNMENT NOR THE CONTRACTOR MAKES ANY WARRANTY, EXPRESS OR IMPLIED, OR ASSUMES ANY LIABILITY FOR THE USE OF CODE WITHIN THIS MANUSCRIPT.

## Author contributions statement

S.D., J.C.B, N.A.K, A.C.J, and B.H.M. conceived the study; K.C, L.C., Y.L.H., S.D., J.D.C., and B.M. implemented data wrangling; S.D. and B.H.M. did most and model development and subsequent analyses; S.D., T.B., R.M.R, N.H., and B.H.M. performed extensive model validation and the interpretation of the modeling results. K.G., J.P.T, J.C.B., N.K. served as the subject matter expert (SME), and together with S.D., and B.H.M. mapped the modeling results to the clinical concepts; S.D. and B.H.M. wrote the manuscript; and all authors reviewed the manuscript.

## Data availability

The data that support the findings of this study are available from Veterans Health Administration (VHA), but restrictions apply to the availability of these data, which were used under license for the current study, and so are not publicly available. Data are however available from the corresponding author upon reasonable request and with permission of the VA.

## Code availability

The overall framework code currently resides in a secure enclave that hosts personal healthcare information (PHI). The relevant portions of the code will be made available by the corresponding author upon reasonable request.

## Glossary

AUPRC: area under the precision-recall curve
AUROC: area under the ROC curve
CCI: Charlson Comorbidity Index
CDC: Center for Disease Control
CDW: corporate data warehouse
CPT: Current Procedural Terminology
CVD: cardiovascular diseases
EMR: electronic medical record
ICD: International Classification of Diseases
ML: machine learning
MVP: Million Veteran Program
NDI: National Death Index
ORD: Office of Research and Development
PC: principal component
PCA: principal component analysis
PHI: personal healthcare information
RCT: randomized controlled trial
RNN: recurrent neural network
SME: subject matter expert
VA: US Department of Veterans Affairs
VACS: Veterans Aging Cohort Study
VHA: Veterans Health Administration

