## SupplementaryInformation for "Data-driven Prediction of Fifteen-Year All-Cause Mortality among 2.3 Million Individuals in the VA"

### SI-1 Patient Characteristics

**Table S1.** Selected characteristics (columns) of groups of patients (rows) selected by demographics, presence of specific medical conditions, and administration of preventative medical procedures. For all groups of patients, the first column describes the selective attribute, together with the specific International Classification of Diseases (ICD)-9 or Current Procedural Terminology (CPT) codes used to define the group. The second provides the total number of patients meeting that criteria together with the percentage of the 2,322,813 total patients in that group. For the *Demographics* groups, the next three columns provide the percentage of each group that are female, Black, and Hispanic. For the *Conditions* and *Procedures* groups, these columns provide the percentage of Females, Blacks, and Hispanics reporting with each condition or procedure reported. For all rows, the last four columns provide the percentage of each group who died, the median age of each group at the times of prediction, the median age at death for those who died, and the median calendar date of death, for those who died.

| **Demographics** | **Total (%)** | **Female** | **Black** | **Hispanic** | **Died** | $Age_{Predict}$ | $Age_{Death}$ | $t_{Death}$ |
|---|---|---|---|---|---|---|---|---|
| All patients | 2,322,813 (100) | 7.89% | 16.1% | 4.70% | 47.6% | 60.7 | 68.75 | 2013.3 |
| Oldest quartile | 580,672 (25.0) | 1.55% | 7.59% | 3.97% | 82.4% | 76.5 | 83.80 | 2012.5 |
| Youngest quartile | 580,693 (25.0) | 18.8% | 27.8% | 5.28% | 17.2% | 47.2 | 56.36 | 2014.4 |
| Female | 183,170 (7.89) | 100% | 16.8% | 2.59% | 16.0% | 50.9 | 60.32 | 2014.7 |
| Black | 373,163 (16.1) | 8.24% | 100% | 1.67% | 36.7% | 55.0 | 63.45 | 2013.7 |
| Hispanic | 109,257 (4.70) | 2.59% | 5.70% | 100% | 38.0% | 59.1 | 67.77 | 2013.9 |
| **Conditions** | | | | | | | | |
| Hypertension (401.9) | 1,320,857 (56.0) | 27.0% | 58.8% | 57.6% | 58.9% | 66.5 | 74.27 | 2013.0 |
| Hyperlipidemia (272.4) | 996,226 (42.9) | 20.3% | 30.6% | 41.5% | 55.3% | 66.2 | 74.31 | 2013.3 |
| Diabetes (250.x) | 557,580 (24.0) | 8.94% | 25.1% | 32.3% | 66.3% | 66.8 | 74.16 | 2012.6 |
| Atherosclerosis (414.0x) | 467,418 (20.1) | 4.15% | 11.7% | 13.5% | 72.4% | 71.3 | 78.12 | 2012.3 |
| COPD (496.) | 341,145 (14.7) | 5.55% | 10.1% | 8.47% | 74.1% | 67.7 | 73.78 | 2011.3 |
| PVD (443.9) | 158,502 (6.82) | 1.63% | 5.53% | 6.72% | 80.7% | 70.9 | 76.91 | 2011.2 |
| Heart failure (428.x) | 142,621 (6.14) | 1.53% | 5.43% | 4.76% | 85.8% | 71.2 | 76.28 | 2010.3 |
| Stroke (43[0-4,8].x) | 121,504 (5.23) | 1.48% | 3.84% | 4.26% | 79.9% | 71.5 | 77.72 | 2011.4 |
| Chronic kidney disease (585.) | 74,222 (3.20) | 0.63% | 4.84% | 3.21% | 83.1% | 70.8 | 75.91 | 2010.4 |
| Colon cancer (153.9 ) | 25,157 (1.08) | 0.32% | 0.93 % | 1.13% | 74.0 % | 72.1 | 78.35 | 2011.5 |
| Prostate cancer (185.) | 121,846 (5.25) | 0.13% | 5.79% | 4.96% | 72.4% | 73.3 | 80.46 | 2012.4 |
| Lung cancer (162.9) | 18,942 (0.82) | 0.23% | 0.71% | 0.41% | 87.3% | 70.2 | 73.39 | 2008.4 |
| Breast cancer (174.9) | 4,023 (0.17) | 1.80% | 0.14% | 0.14% | 47.1% | 59.9 | 66.92 | 2012.3 |
| Depression (311.,296.[23]x) | 521,560 (22.5) | 24.6% | 23.6% | 26.7% | 48.0% | 57.5 | 65.02 | 2012.8 |
| Schizophrenia (295.x) | 97,781 (4.21) | 3.10% | 7.75% | 4.97% | 49.7% | 55.5 | 63.01 | 2012.8 |
| Tobacco use disorder (305.1) | 487,167 (21.0) | 14.0% | 24.9% | 15.7% | 53.6% | 57.5 | 64.72 | 2012.4 |
| Alcoholism (305.0, 303.90) | 222,594 (9.58) | 3.48% | 17.9% | 11.2% | 50.7% | 55.5 | 62.60 | 2012.4 |
| Opioid use disorder (304.00) | 26,665 (1.15) | 0.51% | 3.11% | 1.69% | 49.8% | 52.9 | 60.29 | 2012.7 |
| Homelessness (V60.0) | 85,500 (3.68) | 1.97% | 10.1% | 3.23% | 44.8% | 52.2 | 59.92 | 2013.0 |
| Parkinson's disease (332.x) | 21,767 (0.94) | 0.21% | 0.39% | 0.93% | 86.6% | 73.8 | 78.91 | 2010.4 |
| Alzheimer's disease (331.0) | 11,567 (0.50) | 0.14% | 0.33% | 0.78% | 90.2% | 76.1 | 80.07 | 2009.3 |
| **Procedures** | | | | | | | | |
| Immunization (90471) | 1,113,375 (47.9) | 40.0% | 41.3% | 55.1% | 52.5% | 64.8 | 72.28 | 2012.8 |
| Assay of PSA (84153) | 1,541,358 (66.4) | 1.98% | 64.6% | 72.5% | 52.7% | 64.2 | 71.85 | 2012.9 |
| Occult blood feces (82270) | 1,001,597 (43.1) | 24.6% | 37.3% | 44.1% | 54.9% | 67.0 | 74.57 | 2012.9 |
| Lipid panel (80061) | 1,472,622 (63.4) | 46.6% | 60.4% | 42.5% | 50.2% | 62.7 | 70.34 | 2012.9 |

The prevalence of a variety of medical conditions and common procedures is given in column 2 of Table S1 for three of the most common categories of mortality—cardiovascular disease, cancer, and mental health, as well for four procedures that are representative of preventative medicine. Cardiovascular disease is quite common, with 56% of patients diagnosed with hypertension, and $> 20\%$ each for diabetes and atherosclerosis. Conditions also vary significantly by sex, race, and ethnicity. Hypertension, for example, has less than half the prevalence among females than overall, while diabetes is 33% higher among Hispanics and Blacks than overall. Similarly, the age at diagnosis ranges from 66 for hypertension, hyperlipidemia, and diabetes, up to 71 for stroke and heart failure, while the age at death varies from 74 to 78 across the conditions. The year of death, however, ranges only from 2010 to 2013, reflecting the constraints of our study design.

Similarly large variability is seen in the prevalence, age, and mortality rates of four types of cancers shown. Prostate and breast cancers both have the biases according to sex, prostate cancer is found more often in older patients, and lung cancer has a high mortality rate (87%). As with cardiovascular disease conditions, systematically varying ages of different cohorts of patients makes it difficult to draw conclusions about mortality rates across either conditions or demographic groups by direct inspection of the data.

The third group of conditions shows an even stronger bias effects, as the age range of conditions range from 52 years for homelessness to 76 years for Alzheimer's disease. The 76 year median age of Alzheimer's disease diagnosis happens to match that of the oldest quartile of patients, so it is possible to directly compare probablity of 15-year mortality as being nearly double for the Alzheimer's patients. At the other extreme, patients diagnosed with homelessness or opioid use disorder have a 45% and 50% mortality rate, compared to 17% for the youngest quartile of patients, but the median ages for these groups differ by five years. Similarly, the differences in the fraction of females, Blacks, and Hispanics undergoing each of the procedures shown in Table S1 likely reflects differing age-dependencies recommended for each, convolved with the distinct age distributions of each group.

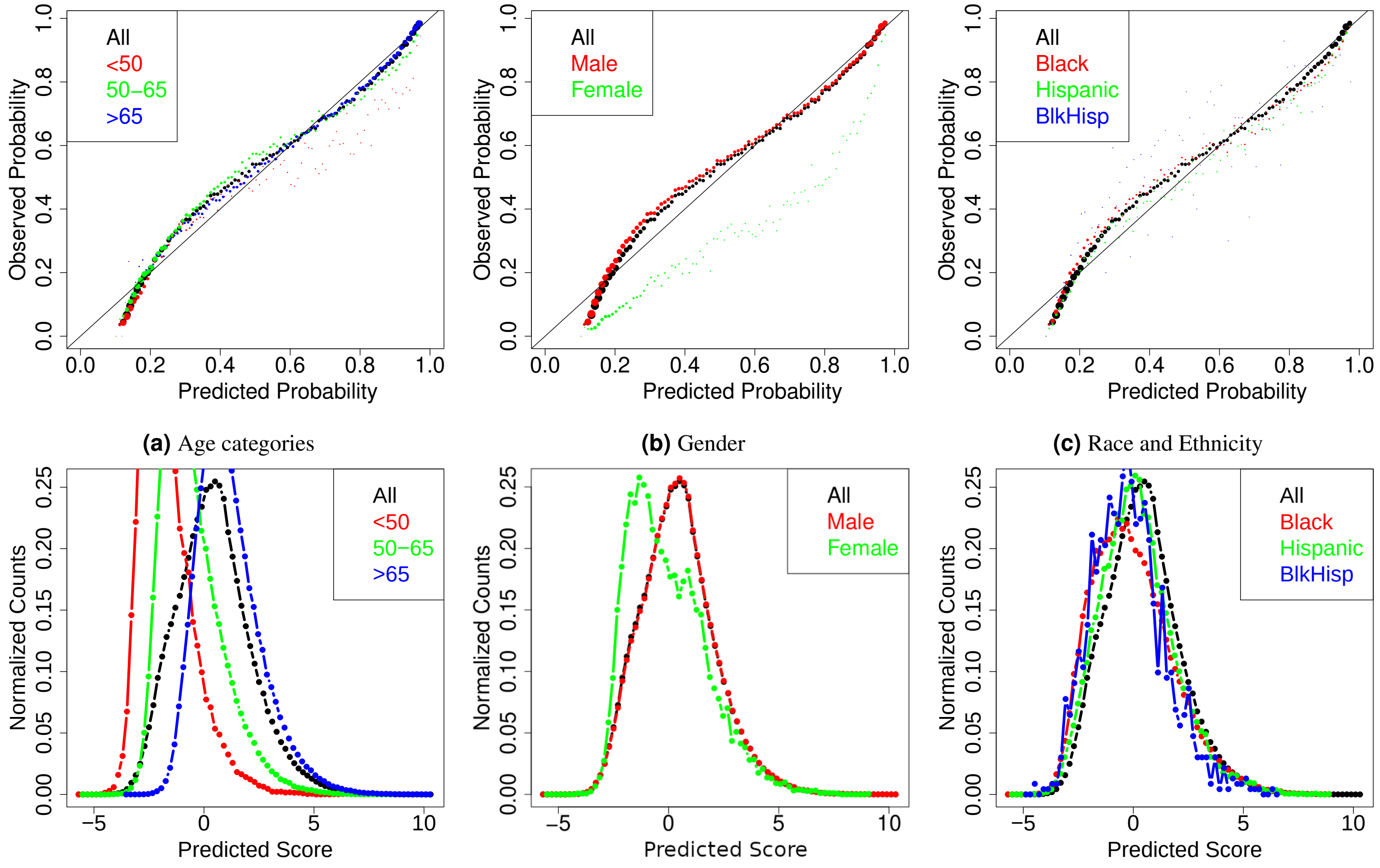


**(a)** Age categories **(b)** Gender **(c)** Race and Ethnicity

**Figure S1.** Calibration plots **(top)** and risk histograms **(bottom)** for our logistic regression model age in the polynomial form, Demo, and all 3000 codes (Dx, CPT, and Rx), across three subgroups defined by age (left), gender (middle), and race / ethnicity (right). The Brier score for this model is 0.155. The predicted score, $S$ (shown as the logit score), is related to the probability, $p$, by the equation, $S = log((p)/(1-p))$. The size (area) of the dots in the top plots is proportional to the number of patients at each score. Subgroups of age and race / ethnicity are well calibrated, while the model overestimates mortality by approximately 30%. The slope and intercept for the calibration are 1.02 and 0.005.

### SI-2 Case Studies from the principal component analysis (PCA) of medical codes

**Cardiovascular-related codes** group towards the bottom of the Figure S3). We see the five largest, common, diagnosis codes of this ICD category to the right of this figure – *Essential hypertension (401.9), Hyperlipidemia (272.4), Uncomplicated diabetes mellitus (250.00), Ischemic heart disease (414.4), and Chronic airway obstruction (496.).* Although these diagnoses are spread across three ICD-9 categories, they are well-known to co-occur and contribute to one-another's disease severity and progression. Four other codes are depicted in large, red symbols, indicating both a high prevalence and that they are relatively strong predictors of mortality. *Hyperlipidemia (272.4)*, however, appears green, as protective in the model. Immediately adjacent to the Dx code for Hyperlipidemia is the symbol for simvastatin prescription, also large and green. Taken together, these may imply that high cholesterol can be readily diagnosed and effectively treated, while the other CVD-related conditions are more complex to manage.

Other drugs in the vicinity of these five diagnoses include lisinopril, an ACE Inhibitor for hypertension; aspirin, to interfere with blood clotting; metoprolol, a beta blocker; nitroglycerin, a vasodilator; furosemide, a diuretic; and metformin, insulin, and glyburide, all commonly used to treat diabetes. Roughly speaking, the PCA is able to successfully associate drug treatments with their associated diagnoses, subject to the condition that more commonly used codes (of whatever type) will be to the right of less commonly used ones. Thus, we can see that simvastatin is of nearly identical prevalence to hyperlipidemia, while lisinopril is taken by only about 2/3 of the patients diagnosed with Hypertension. Similarly, *Ischemic heart disease* is much less common than either hypertension or hyperlipidemia.

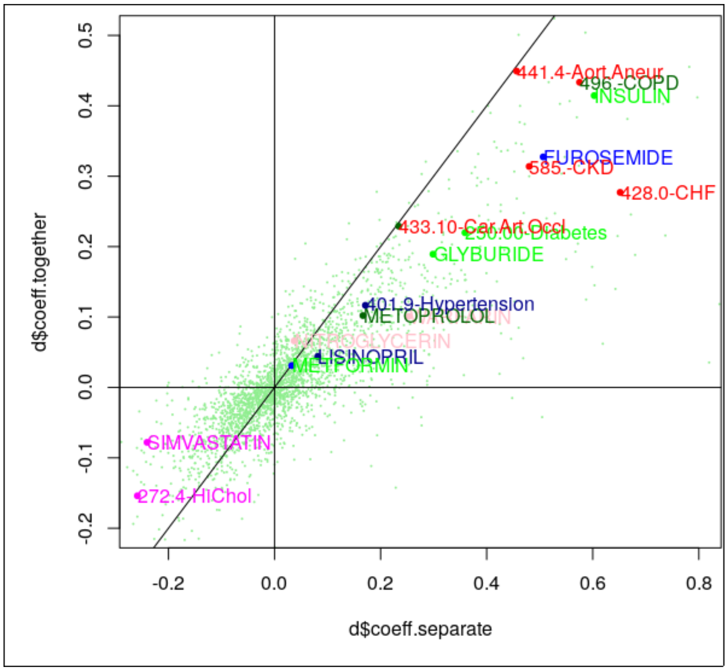


**Figure S2.** Plot of coefficients for models computed with Dx, Rx, and CPT codes included together vs. coefficients for models computed separately. Matching colors pairs of diagnoses/treatments.

Procedure codes can be understood in a similar way. Hemoglobin A1C tests and lipid panels are commonly ordered, among the same patients who are getting other screening tests, such as PSA levels checked, occult blood feces tests, or TSH assays. While most screening tests are protective in predicting all-cause mortality, A1C tests are predictive of mortality, perhaps because the test is ordered only when significant risk of diabetes is present. Another protective procedure is the cardiac stress test, appearing near to *Chronic airway obstruction*.

As we move further to the left in Figure S3, we can see more of the complications of diabetes (retinopathy, renal failure, and peripheral neuropathy) and heart disease (433.10, *Occlusion of the carotid artery*, 585., *Chronic kidney disease*, 428.0, *Congestive heart failure*, and 441.4, *Aortic aneurysm*). Also evident are the CPT codes and drugs associated with treating these conditions, such as those required for heart imaging or revascularization.

The relative placement of the codes vertically is influenced by the comorbidities (and other drugs and procedures) associated with each patient. This is the effect that places mental health diagnoses at the top of Figure S3 while the CVD codes are at the bottom. In a similar way, diabetes conditions are systematically above the CVD-related conditions. For individual treatments, the vertical placement of the code can sometimes indicate prescribing practices dependent on comorbidities.

Similarly, **Mental health-related codes** group towards the top of Figure S3 with five relatively common diagnosis codes predictive of mortality - *depressive disorders (311.x), tobacco abuse disorder (305.1) alcohol dependence syndrome (303.9), alcohol abuse (305.00), homelessness (V60.0)* towards the right of the figure. Interestingly, *major depressive disorder, recurrent type (296.3)*, and *post traumatic stress disorder (309.81)* appear to be protective in the model. Prescriptions of acetaminophen together with codeine or oxycodone, trazodone, gabapentin, sertaline, and hydroxyzine are the most common predictors of mortality while ibuprofen is protective in the model. Procedure codes, understandably, seem to have the biggest effect in mental health related mortality. Psychiatric interview, psychiatric office visit for 20-30 minutes, etc. that are suggestive of a problem being noted appear to be predictive of mortality. However, CPT codes suggestive of therapeutic measures like psychiatric treatment related procedures like psychiatric office visit 45-50 minutes, group psychotherapy, pyschology evaluation of records, and preventive counselling group are all protective.

### SI-3 Clusters of interacting codes

Figure S4 shows three other important clusters identified with the k-means clustering shown in the 3a. We observe interesting patters in the signs of coefficients and whether they change when all three types of medical codes are considered together. An example of this can be seen in the *diabetes* cluster S4a. While the initial diagnosis of uncomplicated diabetes is a significant predictor of mortality, especially with the three versions of uncomplicated diabetes are added together, the relatively low value from 250.50 (diabetes with ophthmalic complications) presumably overlaps with the much larger coefficient from 362.01 (background diabetic retinopathy). The largest single predictor, insulin, is presumably indicating disease severity, as only the more seriously ill diabetic patients are placed on insulin. Two classes of medications, oral sensitizers and ACE inhibitors are represented by three drugs each, with the oral sensitizers highly predictive of mortality and the ACE inhibitors being protective. It is difficult to guess from simple inspection why this difference, but the difference between dark and light bars indicates interactions between the treatments and diagnosis codes.

In addition to this diagnosis-linked indication, 3a shows a graphical representation of the influence of patient behavior (e.g., attending to one's own hygiene) together with externalities (access to housing which in turn in the U.S. typically equates to access to plumbing

**Figure S3.** Multimodal principal component analysis (PCA) plot including all 3000 medical codes, combining results from two calculations: i) the x-y projections are the PCs from the PCA calculation applied to the cosine distance matrix, and ii) the predictive / protective weights are the coefficients from the logistic regression calculation which takes into account the mortality information. The symbol colors represent the sign of the coefficients – red shows predictive and green shows protective of mortality, and the symbol size represent the amplitude of the coefficient. Different symbols are used for the three types of medical codes: '+' for Dx, '×' for CPT, and '◇' for Rx codes.

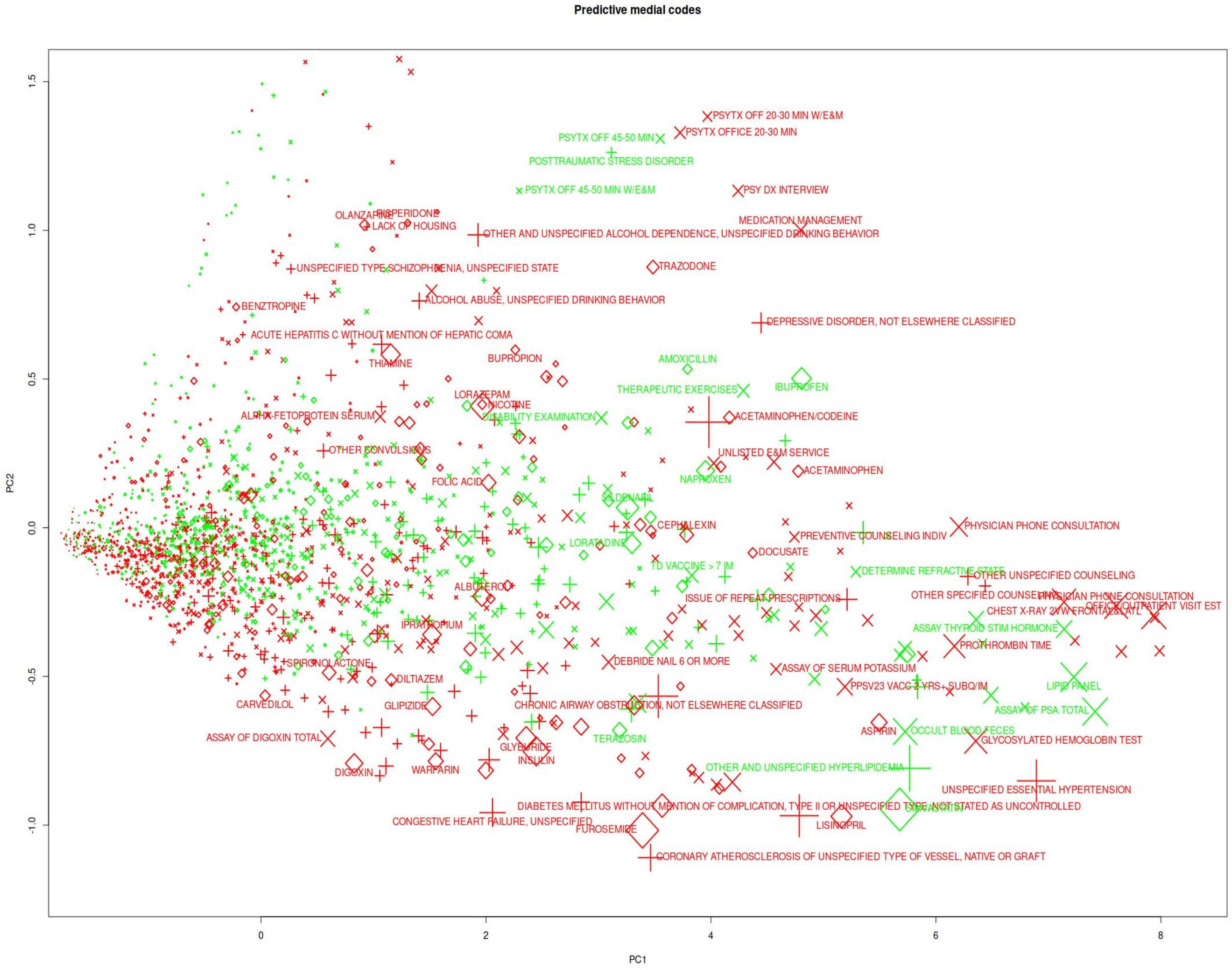


and sewage). In contrast to the diagnosis-related concern for the integrity of the peripheral tissue in the diabetes and PVD case, the influences of behavior and externalities apply across other diagnoses. For instance, a depressed patient with vegetative signs with access to a shower may not attend to hygiene including nail care and may require medical professional intervention. The picture is confounded by the potential for bi-directional causation (for clarity, not drawn): a patient who is unable to provide for her own hygiene and nail care, whether due to an externality such as homelessness or a condition such as arthritis that renders the patient physically unable, may become depressed.

Prima facia the significance to mortality prediction of a given cluster, "nails" for example, may not be intuitively obvious. Persons with Diabetes (either type I or type II) and persons with peripheral vascular disease (PVD) may receive nail care from a medical professional due to the high risk of infection, impaired wound healing, and other medical complications associated with any accidental penetration of the skin, however small.

A somewhat similar narrative with a notable distinction applies to the dental cluster. The distinction is that the dental cluster occupies the dark circle of the center despite dental services being limited to patients identified as high-risk related to financial status. That constraint is muted by broad cross correlation to both the right and left of Figure 3a. And once again, bi-directional influence is plausible. For example, the bacteria associated with poor dental care correlate with cardiac valve disease and myocarditis. In the reverse influence direction, mirroring the previous example, patients who are physically debilitated with cardiac disease (the right of the figure) and/or suffer from mental illness (the left of the figure) may not attend to oral hygiene and require medical professional intervention. The two examples illustrate cross correlation as well as the possibility of influence in either direction. In this case, both are protective, and the coefficients when modeled together add to the value of either coefficient when modeled separately. Second, the treatments can indicate differing levels of disease severity, such as is the case with metformin, glyburide, and insulin, each used to treat 250.00, uncomplicated diabetes. While there are alternative codes (250.1 through 250.9), these codes indicate the types of complications, and do not map well to the type of medication prescribed. Similarly, while some procedures are commonly ordered as screens or preventative medicine, others,

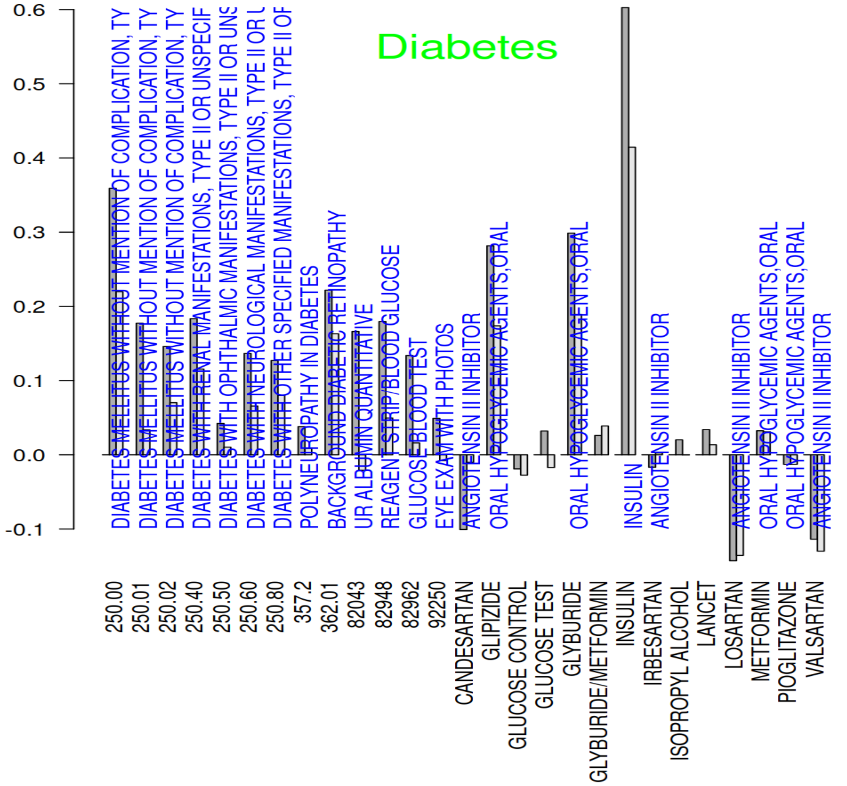


(a)

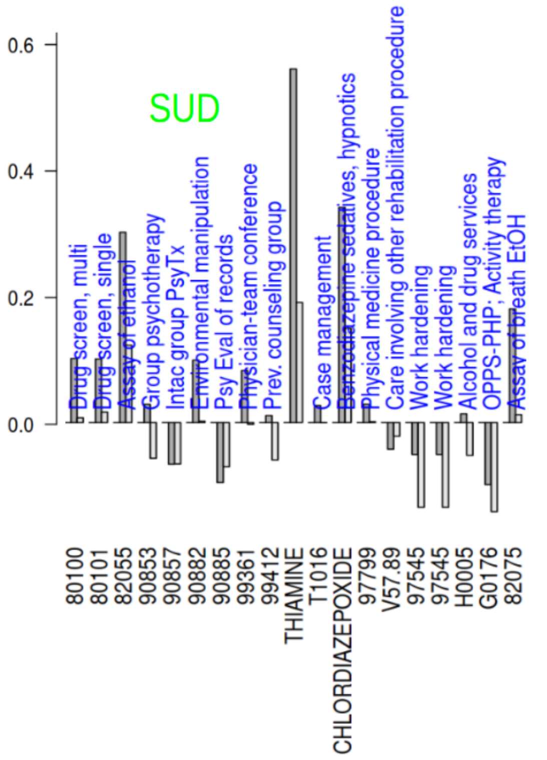


(b)

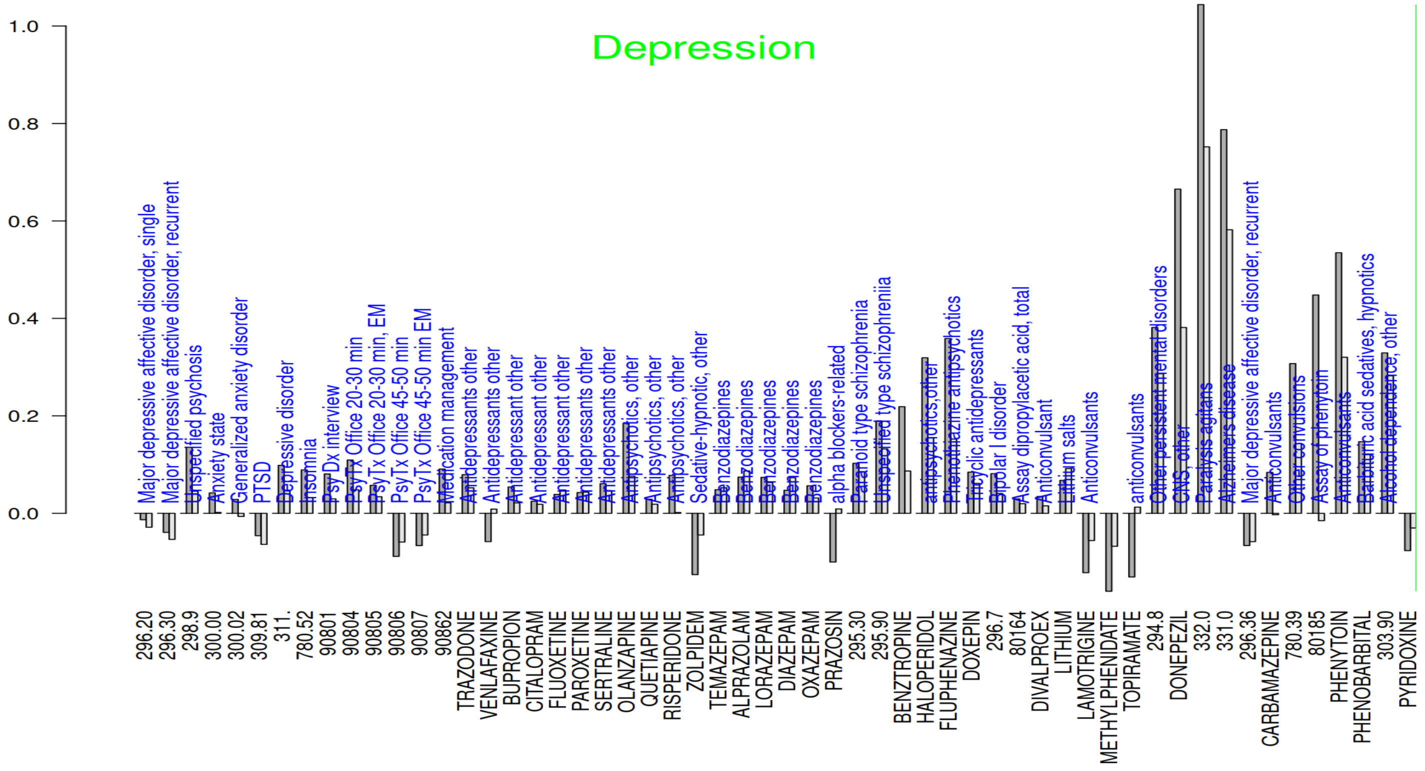


(c)

**Figure S4.** Coefficients of medical codes from the logistic regression models in the **(a) diabetes, (b.) substance use disorder, and (c) depression** clusters taken from Figure 3a. The code is listed at the bottom, while a description is provided over the bars, on the top. Two bars are shown for each code. Gray bars, typically longer, show the model coefficients for the Dx, CPT, or Rx codes from the three models where single type of codes are used as predictors. Clear bars show the coefficients from the single model when all three types of codes are combined as predictors. Positive coefficients extend above and negative below the horizontal line.

such as (HbA1c, EKGs, CT scans, or heart imaging) are ordered only upon suspicion of particular conditions, and also are indicative of disease severity.

For completeness, we provide all the coefficients for the four logistic regression models (Dx-based, CPT-based, Rx-based, and all codes-based), as well as the principal components (PCs) from our principal component analysis (PCA) and the cluster memberships from the K-means clustering in Supplementary File – coefficients-pc-clusters.xlsx

**Table S2.** Impact of randomizing MH and CVD codes on model performance.

| Data Subset | # Individuals | Metric | Original | Randomized MH | Randomized CVD | Randomized MH+CVD |
|---|---|---|---|---|---|---|
| **Entire Test Data** | 1,143,479 | AUROC | 0.849 | 0.834 | 0.831 | 0.814 |
| | | AUPRC | 0.829 | 0.814 | 0.803 | 0.785 |
| **Low Confidence samples (logit $<$ -1)** | 427,485 | AUROC | 0.705 | 0.668 | 0.673 | 0.640 |
| | | AUPRC | 0.325 | 0.286 | 0.285 | 0.263 |

### Glossary

**CPT** Current Procedural Terminology
**ICD** International Classification of Diseases
**PC** principal component
**PCA** principal component analysis
